# Optimizing Temporal Windows for Wearable-Augmented Post-Discharge Risk Prediction: A Methods Study

**DOI:** 10.64898/2026.01.21.26344487

**Authors:** Eric Bressman, Sae-Hwan Park, S. Ryan Greysen, Jinbo Chen

## Abstract

**Objective:** To identify optimal modeling parameters for dynamically predicting hospital readmission risk using post-discharge step-count data from remote monitoring devices.

**Methods:** We combined data from two clinical studies that collected wearable or smartphone-based activity data for up to 6 months after hospital discharge. Analyses were limited to older adults (≥55 years). We constructed a patient-day dataset incorporating static demographic and clinical variables and dynamic activity features aggregated over retrospective windows of 3, 5, 7, or 10 days. Models predicted a composite outcome of readmission or death over prospective horizons of 3, 5, 7, or 10 days, within follow-up periods of 30-180 days. Logistic regression and LightGBM models were trained using 5-fold cross-validation on an 80:20 patient-level split.

**Results:** Among 215 participants, LightGBM outperformed logistic regression across all configurations (mean AUC 0.82 vs 0.76). Performance improved with longer prospective horizons but was largely insensitive to retrospective window length. The LightGBM model was well-calibrated (Hosmer-Lemeshow χ^2^ = 2.46, p = 0.96), whereas logistic regression showed miscalibration (χ^2^ = 51.8, p < 0.001). In feature-importance analyses, LightGBM ranked static (length of stay, vitals, BMI) and dynamic (recent steps, distance) features highly, whereas logistic regression emphasized activity-based variables.

**Discussion:** Prediction performance was impacted by horizon length and training window, with minimal effect of retrospective window. LightGBM achieved higher discrimination and better calibration, supporting flexible, non-parametric methods for post-discharge risk prediction.

**Conclusion:** Post-discharge activity data enhance readmission-risk prediction. Selecting practical temporal windows and appropriate model types can improve accuracy and calibration in wearable-augmented risk models.

## Background

Nearly 17% of older adults discharged from the hospital are readmitted within 30 days, with many of these readmissions considered preventable.^1,2^ Successful transitional care interventions tend to be operationally intensive and difficult to sustain,^3,4^ suggesting the value of an approach targeted toward those with the highest need.^5,6^

Models to predict which patients will be readmitted to the hospital perform poorly, however.^7,8^ In general these models rely on a limited number of predictors and data available at the time of discharge. More recent uses of machine learning (ML) have greatly improved readmission risk prediction, as ML allows for consideration of a large number of predictors and offers greater flexibility in identifying non-linear relationships between predictors and outcome.^9,10^ The addition of remote monitoring data streams – which provide a window into how patients are doing after discharge – improves risk prediction even further.^11,12^ Remotely monitored activity data, in particular, via wearable devices or smartphone apps, has been shown to greatly improve risk prediction.^13^

Little is known, however, about optimal modeling parameters when using step count data to dynamically predict risk. Prior studies have generally relied on intuition to make decisions about 1) engineering of step count features and 2) optimal prediction windows. The former is important because it may have implications for both model performance and the amount of daily activity data that needs to be retained to run the model prospectively. One study, for instance, relied on minute-level data,^14^ which is impractical given how much missingness is typically present in real-world activity-monitoring programs.^15^ The latter has implications for how these models are implemented in clinical settings and how feasibly the risk outputs can be acted upon.

In this methodologic study, we use a novel dataset combining clinical and activity data from older adults after discharge to explore the optimal modeling parameters for dynamically predicting readmission risk. We compare lengths of retrospective step count windows (the number of days of past activity data used), prospective prediction windows (over what time period the model forecasts risk), and training windows (the length of time post-discharge for which data is used). To evaluate the robustness of our findings, we compare models trained with both a parametric method (logistic regression) and a non-parametric method (gradient-boosted trees).

## Methods

### Patients and Data Sources

We used data from patients enrolled in 2 clinical studies at Penn Medicine which ran between January 2017 and December 2019 (PREDICT and MOVE IT).^13,16^ In both studies, patients used either a wearable device or a smartphone application to track activity data for at least 3 months after hospital discharge. Demographic and clinical data from these studies came from the electronic health record (EHR) via the Clarity database (Epic’s reporting database). Additional sociodemographic data for patients in both studies were collected via enrollment surveys administered at the start of the studies. Finally, data transmitted from remote monitoring devices were stored in Way To Health, a cloud-based research technology platform at the University of Pennsylvania used to support digital and behavioral health clinical studies.^17^ The study was approved by the University of Pennsylvania Institutional Review Board.

### Datasets

#### PREDICT

Prediction using a Randomized Evaluation of Data collection Integrated through Connected Technologies (PREDICT) was a 2-arm clinical trial which randomized patients to use of either a smartphone application or a wearable device to collect mobility data after discharge from the hospital. Patient data collected included static and dynamic covariates. The static covariates included: socio-demographics (age, sex, race/ethnicity, education, marital status, annual household income), and clinical data (Charlson Comorbidity Index, body mass index, vitals [the values closest to discharge for temperature, heart rate, systolic and diastolic blood pressures, oxygen saturation, and respirations per minute], and hospital length of stay). The dynamic covariates came from wearable devices: activity data (daily measures of step counts, distance, calories burned [active and total], and minutes of activity [soft, moderate, and intense]) and sleep data (minutes of sleep [total, light, and deep], minutes to fall asleep, and number of times awakened). The devices (smartphone and wearable) generated comparable data for prediction. Outliers (top and bottom 1 percentile) in the activity data were coded as missing. Missingness was addressed through a combination of missing-value indicator variables and multiple imputation by chained equations, using patient demographics, clinical covariates, and time indicators as predictors. The investigators also included measures of missingness itself (e.g., proportion of days with captured data) and lagged weighted averages of activity over the prior 3 days, to capture recent changes in behavior. Full detail on data cleaning and feature engineering is available in the initial study.^13^

#### MOVE IT

Mobility and Outcomes for Validated Evidence–Incentive Trial (MOVE IT) was a 2-arm clinical trial which provided all participants with a wearable device to collect mobility data after hospital discharge, and randomized them to an intervention (a game informed by behavioral economics to incentivize reaching step goals) or control (no such game). Patient data collected included demographics (age, sex, race/ethnicity, insurance type, education, marital status, annual household income), clinical data (Charlson Comorbidity Index, discharge diagnosis, body mass index, hospital length of stay), and remote monitoring data (same data elements as described for PREDICT above). Any data elements that were not collected at the time of the original study were pulled from the Clarity database for the purposes of this analysis. In the original study, step counts <1000 were considered implausible and coded as missing. Missing or implausible values were imputed using multiple imputation with five sets of imputations, and results combined using Rubin’s rules. Full detail on data cleaning and feature engineering is available in the initial study.^16,18^

For the current study, we re-engineered the MOVE IT dataset to support dynamic prediction modeling. We applied an identical preprocessing and feature-generation pipeline to all participants, regardless of study and/or trial arm, ensuring comparability between the PREDICT and MOVE IT datasets.

### Feature Engineering

We combined the PREDICT and MOVE IT data into a single dataset, and then selected out the subset of older adults (age 55+) to create our final training dataset (n = 215). Data was represented at the patient-day level, with each row representing one participant on one day after discharge. Static variables (sociodemographic and clinical data elements) were carried forward unchanged.

For every dynamic signal transmitted by the wearable or smartphone—daily steps, distance, active/total calories, soft/moderate/intense activity minutes, and multiple sleep metrics—we extracted two summary statistics over a retrospective window of *m* days (3, 5, 7 or 10):

- Mean value during retrospective window
- Proportion of days with missing data during retrospective window

Windows were anchored so that features used information from day max(1, *a - m +* 1) through day *a* after discharge (where *a* represents the current day).

### Model Development

#### Prediction framework

At each index day *a*, the model forecasted the composite outcome of unplanned readmission or death occurring over the next *n* days (prospective horizon = 3, 5, 7 or 10) provided *a + n* ≤ *k*, where *k* is the maximum follow-up window examined (30, 60, 90 or 180 days) (Figure 1).

#### Data splitting

Data were stratified at the patient level into 80% training and 20% held-out test sets, ensuring all daily observations for a given individual resided in only one split.

#### Model training

Within the training set, we trained models using both a classical statistical method (multivariate logistic regression) and a machine learning method, gradient boosting (LightGBM). Hyper-parameters for LightGBM (learning rate, maximum depth, number of leaves, L2 penalty and subsampling rates) were tuned inside five-fold cross-validation on the training partition. We trained models using both methods for every combination of (*m, n, k*), where *m* was the retrospective window for looking at dynamic variables, *n* was the prospective window for predicting the outcome, and *k* was the maximum period of time after discharge used for training.

### Statistical Analysis

Model discrimination was quantified with the area under the receiver-operating-characteristic curve (ROC-AUC). We computed ROC-AUCs on the held-out test set for every combination of (*m, n, k*) and for both algorithms. Model calibration was assessed using the Hosmer-Lemeshow goodness-of-fit test, which compares observed and expected event rates across deciles of predicted risk (χ^2^ statistic with 8 degrees of freedom).

To understand how design choices affected performance, we:

- Summarized mean ROC-AUC across all combinations for each algorithm.
- Plotted ROC-AUC against each temporal parameter while holding the others constant.

Significance was defined by two-sided *p* < 0.05. All analyses were conducted using Python 3.13. Data manipulation and statistical analyses were performed using Python packages: Pandas, NumPy, and Statsmodels. Machine learning models were implemented using Scikit-Learn and LightGBM packages.

## Results

Our overall cohort (training and test sets) consisted of 215 individuals, among which the mean (SD) age was 61.3 (5.2); 124 (57.7%) were female; 80 (37.2%) were Black and 125 (58.1%) were White; and 32 (14.9%) experienced a 30-day readmission. The mean (SD) step count was 2684.2 (2497.5) and days with missing data per week was 1.6 (2.5). Characteristics were generally similar across the two source studies, with no significant differences in age, sex, comorbidity, or mean step count (Table 1). Patients from MOVE IT had marginally shorter hospital stays (3.9 vs 7.4 days) and somewhat higher education and income levels, but clinical and activity measures were otherwise comparable.

**Table 1.**
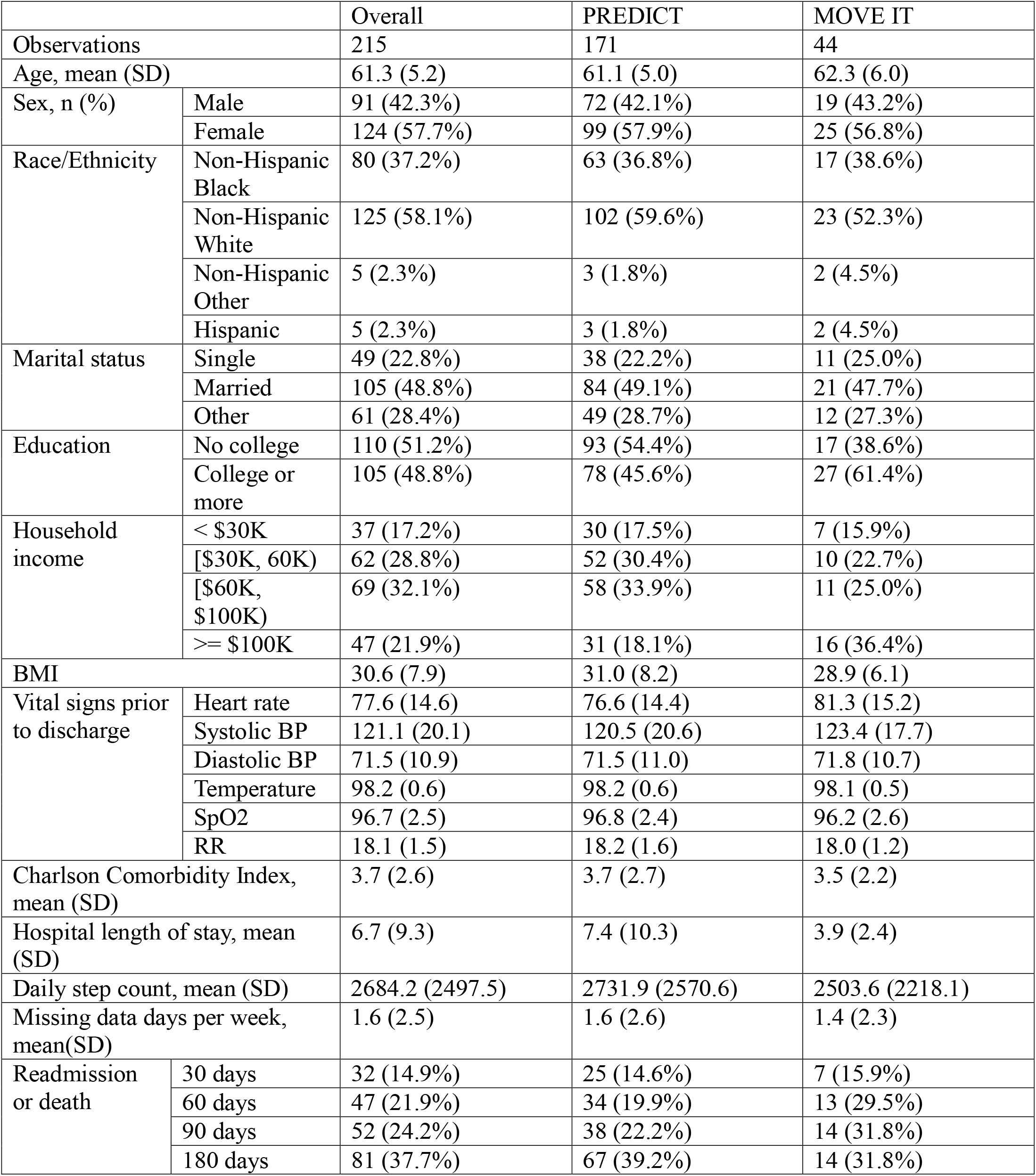
Cohort characteristics.

|  |  | Overall | PREDICT | MOVE IT |
| --- | --- | --- | --- | --- |
| Observations |  | 215 | 171 | 44 |
| Age, mean (SD) |  | 61.3 (5.2) | 61.1 (5.0) | 62.3 (6.0) |
| Sex, n (%) | Male | 91 (42.3%) | 72 (42.1%) | 19 (43.2%) |
|  | Female | 124 (57.7%) | 99 (57.9%) | 25 (56.8%) |
| Race/Ethnicity | Non-Hispanic Black | 80 (37.2%) | 63 (36.8%) | 17 (38.6%) |
|  | Non-Hispanic White | 125 (58.1%) | 102 (59.6%) | 23 (52.3%) |
|  | Non-Hispanic Other | 5 (2.3%) | 3 (1.8%) | 2 (4.5%) |
|  | Hispanic | 5 (2.3%) | 3 (1.8%) | 2 (4.5%) |
| Marital status | Single | 49 (22.8%) | 38 (22.2%) | 11 (25.0%) |
|  | Married | 105 (48.8%) | 84 (49.1%) | 21 (47.7%) |
|  | Other | 61 (28.4%) | 49 (28.7%) | 12 (27.3%) |
| Education | No college | 110 (51.2%) | 93 (54.4%) | 17 (38.6%) |
|  | College or more | 105 (48.8%) | 78 (45.6%) | 27 (61.4%) |
| Household income | < \$30K | 37 (17.2%) | 30 (17.5%) | 7 (15.9%) |
| | [\$30K, 60K) | 62 (28.8%) | 52 (30.4%) | 10 (22.7%) |
| | [\$60K, \$100K) | 69 (32.1%) | 58 (33.9%) | 11 (25.0%) |
| | >= \$100K | 47 (21.9%) | 31 (18.1%) | 16 (36.4%) |
| BMI |  | 30.6 (7.9) | 31.0 (8.2) | 28.9 (6.1) |
| Vital signs prior to discharge | Heart rate | 77.6 (14.6) | 76.6 (14.4) | 81.3 (15.2) |
|  | Systolic BP | 121.1 (20.1) | 120.5 (20.6) | 123.4 (17.7) |
|  | Diastolic BP | 71.5 (10.9) | 71.5 (11.0) | 71.8 (10.7) |
|  | Temperature | 98.2 (0.6) | 98.2 (0.6) | 98.1 (0.5) |
|  | SpO2 | 96.7 (2.5) | 96.8 (2.4) | 96.2 (2.6) |
|  | RR | 18.1 (1.5) | 18.2 (1.6) | 18.0 (1.2) |
| Charlson Comorbidity Index, mean (SD) |  | 3.7 (2.6) | 3.7 (2.7) | 3.5 (2.2) |
| Hospital length of stay, mean (SD) |  | 6.7 (9.3) | 7.4 (10.3) | 3.9 (2.4) |
| Daily step count, mean (SD) |  | 2684.2 (2497.5) | 2731.9 (2570.6) | 2503.6 (2218.1) |
| Missing data days per week, mean(SD) |  | 1.6 (2.5) | 1.6 (2.6) | 1.4 (2.3) |
| Readmission or death | 30 days | 32 (14.9%) | 25 (14.6%) | 7 (15.9%) |
|  | 60 days | 47 (21.9%) | 34 (19.9%) | 13 (29.5%) |
|  | 90 days | 52 (24.2%) | 38 (22.2%) | 14 (31.8%) |
|  | 180 days | 81 (37.7%) | 67 (39.2%) | 14 (31.8%) |

### Overall model performance

Across the full grid of temporal configurations, the gradient-boosted tree ensemble (LightGBM) outperformed the baseline logistic model. Test set mean ROC-AUC for LightGBM (across all the experiments) was 0.82 versus 0.76 for logistic regression, and its performance stability was similar (SD = 0.06 vs 0.07).

Calibration results mirrored the discrimination findings. The best LightGBM model showed good calibration by the Hosmer–Lemeshow test (χ^2^ = 2.46, df = 8, *p* = 0.96), supporting the null hypothesis of good fit. In contrast, the best logistic-regression model demonstrated evidence of miscalibration (χ^2^ = 51.8, df = 8, *p* < 0.001), suggesting that the linear specification may over- or under-estimate absolute risk in certain ranges of predicted probability.

### Impact of temporal window parameters

Figure 1 shows discrimination by retrospective window (*m*). Performance was largely insensitive to how many prior days of activity were summarized; only a modest uptick was seen when the window was extended to 7-10 days.

**Figure 1.**
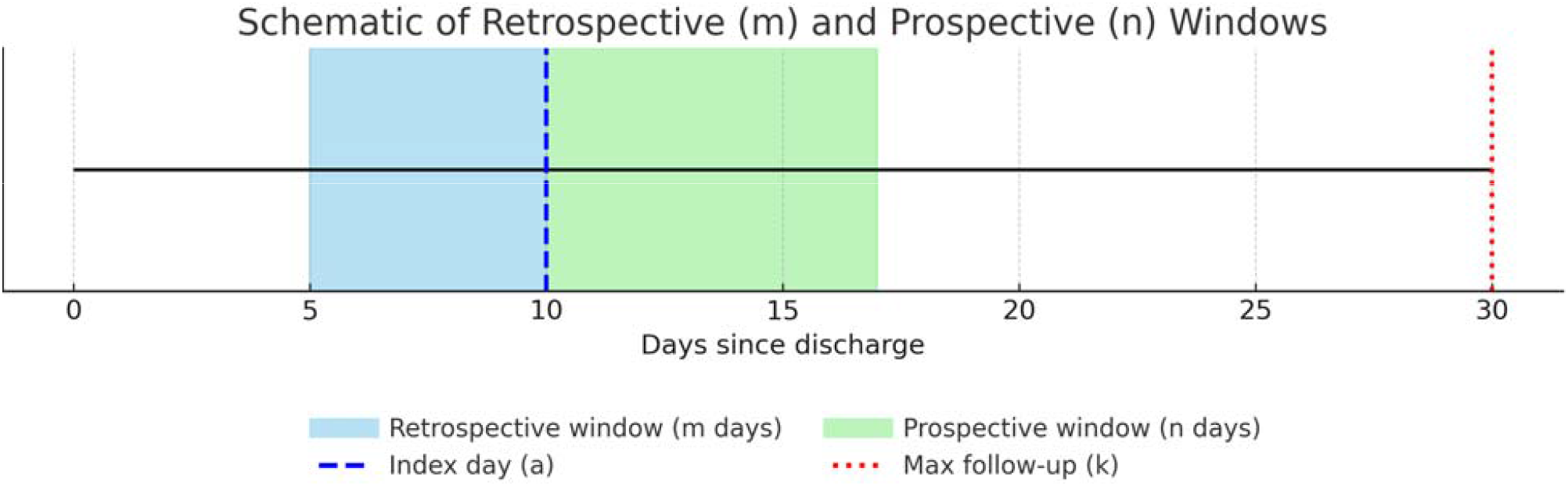

As illustrated in Figure 2, longer prospective prediction horizons (*n*) improved discrimination for both algorithms; a 10-day horizon yielded the highest accuracy (ROC-AUC 0.87 for LightGBM vs 0.79 for logistic regression).

**Figure 2.**
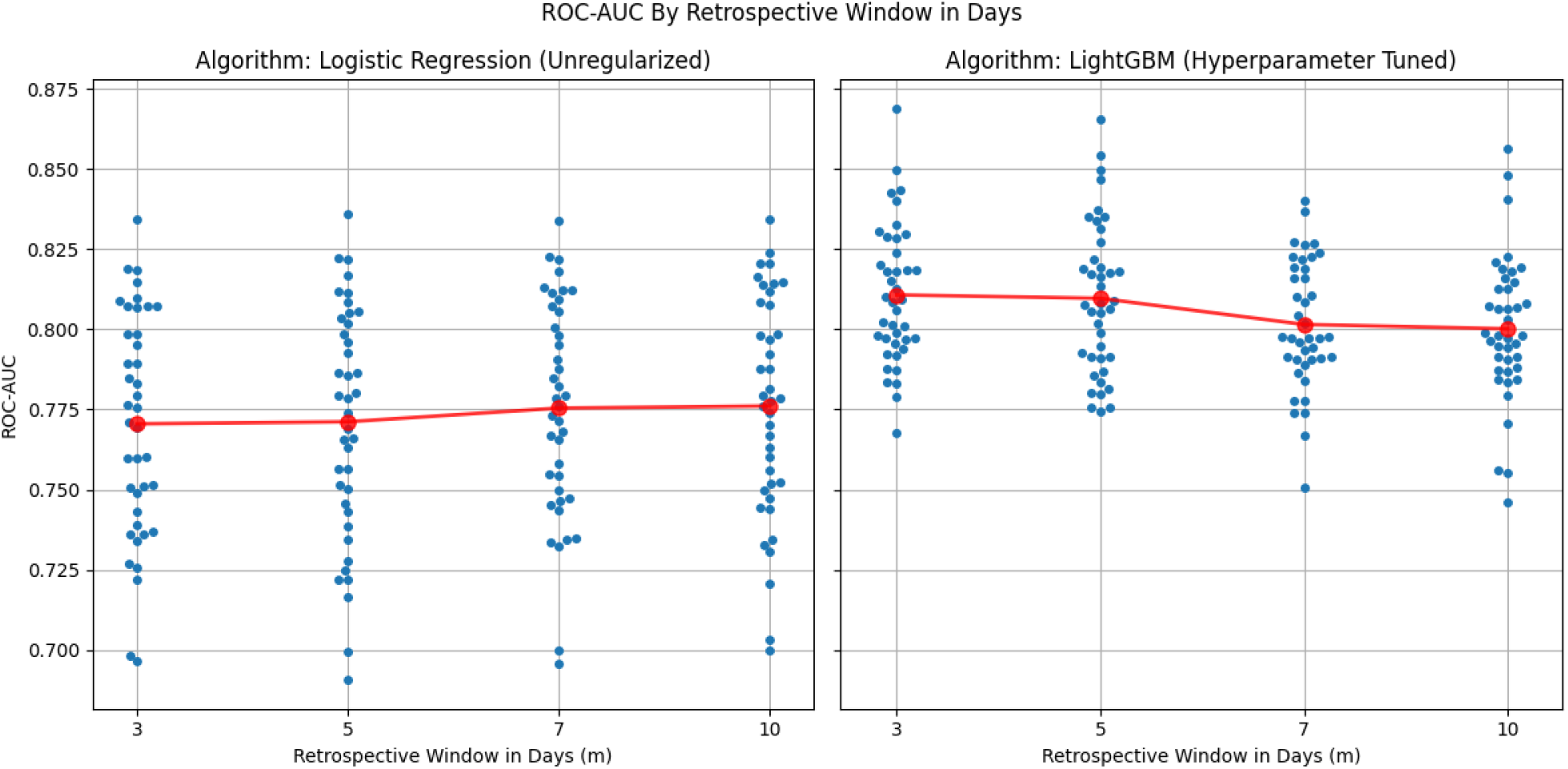
Relationship between ROC-AUC and Respective Window Size by Training Method.

Varying the training window (*k*) (the maximum follow-up period after discharge included in model fitting) produced a non-monotonic pattern (Figure 3). Logistic regression performed best with 90-day windows (mean ROC-AUC 0.81) followed by 30 days, whereas LightGBM peaked when restricted to 30 days (mean ROC-AUC 0.83) followed by 90 days. Discrimination for both models was lowest when the training window was extended out to 180 days after discharge.

**Figure 3.**
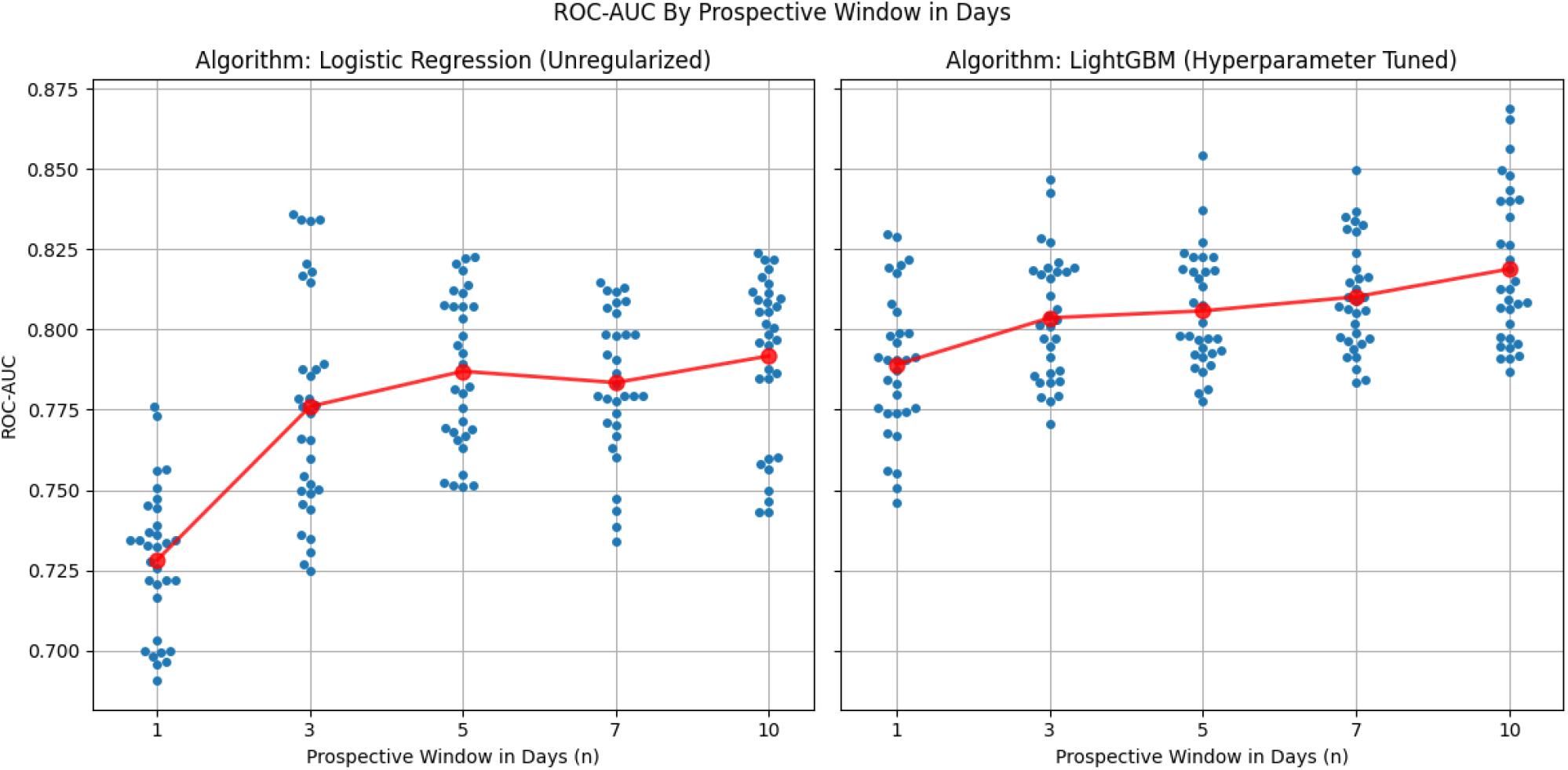
Relationship between ROC-AUC and Prospective Window Size by Training Method.

### Feature importance

Feature importance was examined for the best-performing LightGBM and logistic regression models (Table 2). In the LightGBM model, the top-ranked features by total loss reduction were hospital length of stay, systolic blood pressure, and body mass index. In the logistic regression model, the features with the largest absolute coefficients were mean intense-activity minutes, mean step count, and mean distance.

**Table 2.**
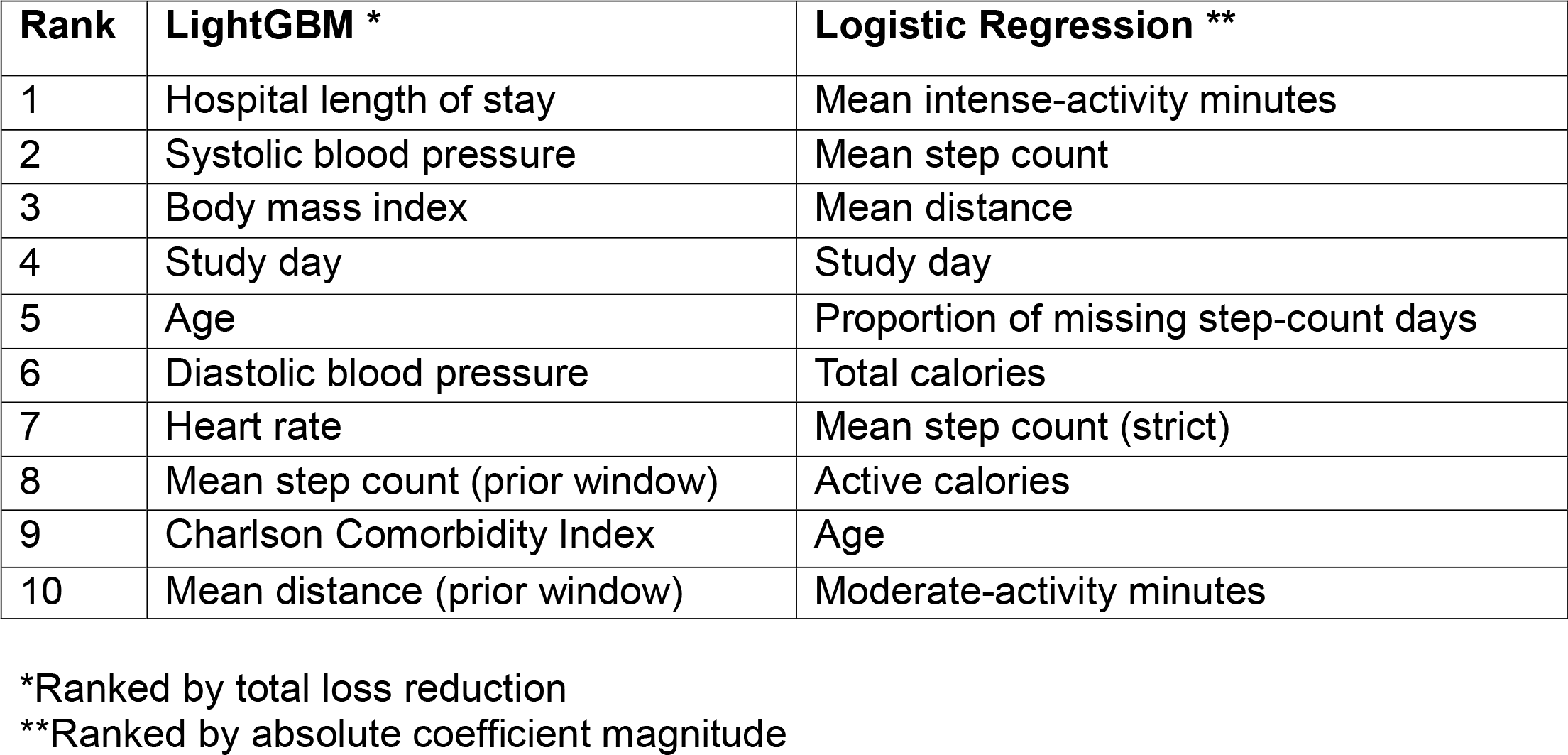
Top 10 features ranked by importance for LightGBM and logistic regression models.

| Rank | LightGBM * | Logistic Regression ** |
| --- | --- | --- |
| 1 | Hospital length of stay | Mean intense-activity minutes |
| 2 | Systolic blood pressure | Mean step count |
| 3 | Body mass index | Mean distance |
| 4 | Study day | Study day |
| 5 | Age | Proportion of missing step-count days |
| 6 | Diastolic blood pressure | Total calories |
| 7 | Heart rate | Mean step count (strict) |
| 8 | Mean step count (prior window) | Active calories |
| 9 | Charlson Comorbidity Index | Age |
| 10 | Mean distance (prior window) | Moderate-activity minutes |

\*Ranked by total loss reduction
\*\*Ranked by absolute coefficient magnitude

## Discussion

In this methods study we systematically varied three temporal design choices in training a dynamic readmission risk model using step count features: (1) the length of the retrospective look-back window for step-count features, (2) the prospective prediction horizon, and (3) the overall training window, using both linear (logistic regression) and non-linear (ensemble trees with gradient-boosting) training methods. Model performance was insensitive to the amount of historical activity data beyond three days, improved monotonically with longer prediction horizons up to 10 days, and declined when the training window exceeded 90 days.

These patterns suggest that (a) the most recent activity captures the salient signal, and (b) near-term risk can be forecast with sufficient lead time for transitional-care teams to intervene. Calibration findings further highlight that the LightGBM model not only achieved higher discrimination but also maintained good alignment between predicted and observed risk, suggesting its suitability for generating actionable probability estimates in clinical use. By contrast, the logistic regression model demonstrated poorer calibration, indicating potential over-or under-estimation of absolute risk despite moderate discrimination.

Our overall discrimination (mean AUC 0.82) exceeds that of many recent machine-learning readmission risk models trained on static data alone (median AUC 0.68, IQR: 0.64-0.76)^19^ and is comparable to the few wearable-augmented studies published to date. Nagarajan et al integrated Fitbit entropy metrics with clinical data and achieved AUC 0.83 for 90-day readmission;^14^ the original PREDICT study, from which a portion of our dataset derived, achieved similar results for the similar age subgroup.^13^ The former study used a multiscale entropy technique for modeling the dynamic data, which allows for characterization of temporal variation including at the intraday level; this is computationally intensive, and also runs the risk of overfitting. The latter study included summary metrics over multiple retrospective windows.

Both studies made predictions at the day level (i.e., what is the likelihood that the patient will be readmitted that day), and did not explore longer prediction horizons. On a practical level, it would be challenging to build a transitional care intervention around same day predictions – staff rarely have the bandwidth to respond so quickly, and it would likely require weekend and off hours staffing. In our analysis, both algorithms performed poorest with same-day predictions, and demonstrated stable to improved performance with longer prediction horizons. Henderson et al looked at varying prediction horizons with a static prediction model (using claims data), but varied the windows over longer windows (14, 30, and 60 days); they found decreasing performance with increasing horizons.^20^ To our knowledge our study is the first to both compare shorter windows and use dynamic features.

Feature □ importance analyses suggested that both static clinical characteristics (e.g., hospital length of stay, vital signs) and recent activity measures contributed meaningfully to prediction. The relative prominence of static features in LightGBM versus dynamic activity variables in logistic regression reflects methodological differences rather than inconsistent signal, underscoring that wearable and clinical data provide complementary information.

Strengths of this study include a large multi-study cohort with day-level activity data; a fully crossed experimental design isolating the effect of each temporal parameter; and rigorous patient-level splits that prevent information leakage. Limitations include that all participants came from a single academic center and consented to participation, potentially limiting generalizability. Second, we focused on tree-based ensembles; deep-learning architectures or survival models might yield further gains. Finally, external validation and impact analyses are still required before clinical deployment. Future work might entail validating the highest performing model in diverse settings; evaluating decision-curve utility to select actionable risk thresholds; and embedding the algorithm in a pragmatic randomized trial of transitional-care interventions.

## Conclusions

Leveraging remotely monitored step count data and machine learning model training methods both meaningfully enhance dynamic readmission risk prediction in older adults. Our work shows that shorter historical windows and moderately long prediction horizons strike the best balance between accuracy and operational feasibility, providing a blueprint for health systems seeking to develop such models to target resource intensive transitional care services to those most likely to benefit.

## Data Availability

N/A

